# Balancing Analgesic Effects and Abuse Liability in Older Adults: A Randomized, Placebo-Controlled Trial of Oral and Vaporized Δ9-Tetrahydrocannabinol

**DOI:** 10.64898/2026.11.02.26367127

**Authors:** Gabriel P. A. Costa, Ralitza Gueorguieva, Matthew M. Riggs, Tyler Dunlap, Mehmet Sofuoglu, Joao P. De Aquino

## Abstract

Cannabis use is rising fastest among older adults in the U.S., and pain is among the most common reasons they use it. No trial, however, has measured the trade-off between the analgesic and abuse-related effects of Δ9-tetrahydrocannabinol (Δ9-THC) in this population. In a randomized, double-blind, placebo-controlled trial, 20 adults aged 65 and older were assigned to oral Δ9-THC (placebo, 5, 10 mg) or vaporized Δ9-THC (placebo, 2, 4 mg) across three eight-hour sessions in counterbalanced order. We assessed laboratory-evoked pain sensitivity using quantitative sensory testing, abuse liability using subjective drug effects and reinforcement value, and also sampled plasma Δ9-THC and 11-OH-THC. Analgesia and abuse liability outcomes were measured repeatedly within each session. On a pain sensitivity index, oral 5 mg relieved pain relative to placebo (p=.040), although the oral arm’s overall dose effect was not significant (p=.108); oral 10 mg and both vaporized doses did not differ from placebo. Subjective effects increased with dose and were greatest after vaporization, whereas reinforcement value increased only after vaporized 4 mg. After oral 5 mg, analgesia exceeded placebo at every assessment (60-360 min), while its pleasurable effects peaked at 150 min. Vaporized Δ9-THC produced pleasurable effects within 30 min but did not significantly change thermal pain sensitivity. While abuse liability tracked Δ9-THC exposure, analgesia showed no significant association, and all adverse events occurred at vaporized 4 mg. These findings suggest dose and route selection may help maximize Δ9-THC analgesia while minimizing abuse liability, a strategy that warrants evaluation in clinical populations.

## 1. Introduction

Chronic pain affects over a third of adults aged 65 and older[33], and many older adults have turned to cannabis to relieve their pain. Older adults are now the fastest-growing group of people who use cannabis: past-year use has risen 20-fold in two decades[22,23,40,47], and more than half of older adults who use cannabis cite pain as a reason[6,44,47,55]. Between 2015 and 2019, the share of older adults who see regular use as a great risk fell from 53% to 43%[21]. Chronic pain is also a risk factor for cannabis use disorder (CUD), and CUD diagnoses are rising fast among older adults[25].

Δ9-tetrahydrocannabinol (Δ9-THC) is a partial agonist at cannabinoid type 1 (CB1) receptors[46] and reliably produces analgesia in preclinical models[24,35,36]. Clinically, however, cannabis and Δ9-THC relieve chronic pain modestly: a minimally important improvement in 10% more patients than placebo[54]. Laboratory findings from quantitative sensory testing (QST), which evokes pain with calibrated stimuli[48], are inconsistent. A systematic review of 39 placebo-controlled QST studies of inhaled cannabis, dronabinol, nabilone, Δ9-THC:CBD formulations, and other cannabinoid agonists, almost all in younger adults, found them too heterogeneous to pool[42]. Controlled studies have evaluated the pharmacokinetics, tolerability, or dementia-related agitation associated with cannabis in older adults, but no studies have evaluated analgesia[2,3].

Aging reshapes both the nociceptive system and the pharmacokinetics and pharmacodynamics of Δ9-THC[15]. Pain thresholds tend to increase with age, whereas pain tolerance changes are more modest[34]. CB1 receptor availability declines with age in preclinical models[19], and in human frontal cortex post mortem, receptor density and function fall by roughly 10% per decade[39]. Emerging data indicate that the cognitive effects of acute Δ9-THC are blunted in older adults[15]. Greater adiposity and reduced hepatic blood flow also prolong exposure to Δ9-THC and its active metabolite 11-OH-THC[14,30,38]. These changes may attenuate, potentiate, or leave unchanged Δ9-THC’s analgesic and abuse-related effects, and not necessarily in the same direction for each[15].

Dose and route can also affect analgesia and abuse liability differently[13]. Δ9-THC’s dose-response relationship is non-linear and often U-shaped[9]: in healthy volunteers, an intermediate dose of smoked cannabis reduced capsaicin-induced pain, whereas a higher dose increased it[53]. In contrast, abuse liability, tends to increase with dose[50] and with the speed at which a drug reaches the brain[1]. Route sets that speed: among older adults, vaporized Δ9-THC produced peak concentrations roughly nine-fold higher than matched oral doses, at 3 minutes rather than 60 to 90 minutes[14]. Taken together, these results suggest the benefit-risk profile of cannabis in older adults could favor lower doses (i.e., greater analgesia and lower abuse liability).

Both outcomes therefore have to be measured together. To identify this trade-off in older adults, we conducted a randomized, double-blind, placebo-controlled trial of oral (5 and 10 mg) and vaporized (2 and 4 mg) Δ9-THC, measuring both outcomes repeatedly in each session. We hypothesized an inverted-U dose-response for analgesia, placing the most favorable benefit-risk balance at the lower dose of each route. We also asked whether analgesia tracks total exposure while abuse liability tracks peak concentration.

## 2. Methods

We prospectively registered this study (ClinicalTrials.gov NCT05906511) and the protocol was approved by the Yale University and VA Connecticut Healthcare System Institutional Review Boards. The study was conducted at the Biological Studies Unit of the VA Connecticut Healthcare System (West Haven, CT). All participants provided written informed consent before any study procedure, and reporting follows the Consolidated Standards of Reporting Trials (CONSORT) 2025 statement[27]. This report covers laboratory-evoked pain and its balance against abuse liability. All other outcomes are reported in the companion manuscript[14].

### 2.1 Study Design

The trial design is described in full in the companion manuscript[14]. Briefly, participants were randomized to one route of Δ9-THC, oral or vaporized, and within that arm completed three eight-hour test sessions, one per dose (oral: placebo, 5 mg, 10 mg; vaporized: placebo, 2 mg, 4 mg). Dose order was randomized and counterbalanced, and sessions were separated by at least 72 hours to mitigate carryover.

### 2.2 Participants

Participants were healthy adults aged 65 years or older who had prior lifetime cannabis exposure and no DSM-5 psychiatric or substance use disorder, other than tobacco use disorder, in the past year. We required lifetime cannabis exposure to enhance the safety and tolerability of the study. Because the present analyses depend on intact nociception, eligibility excluded conditions that alter pain sensation or balance (for example, stroke or peripheral neuropathy identified on a neuro-sensory examination), contraindications to nociceptive stimulation such as untreated hypertension, and regular use of analgesics or of strong CYP2C9, CYP3A4, or UGT1A9 modulators. A negative urine test for THC-COOH (50 ng/mL cutoff) was required for enrollment. Medical exclusions included hepatic enzyme elevation and renal dysfunction.

Psychiatric exclusions included a personal or family history of primary psychotic or mood disorders with psychotic features. Full eligibility criteria are listed in **Supplementary Section S1**.

### 2.3 Study Medication and Administration

Oral Δ9-THC was administered as dronabinol (synthetic Δ9-THC; McKesson Corporation, Irving, TX), an FDA-approved Schedule III oral formulation, in capsules containing 0, 5, or 10 mg. Vaporized Δ9-THC was delivered as pharmaceutical-grade Δ9-THC (99% purity, compounded as a 10 mg/mL ethanolic solution) using the Volcano Hybrid® vaporizer (Storz & Bickel, Tuttlingen, Germany)[32] with a standardized paced-puffing procedure[8,17] described in the companion manuscript. Active vaporized doses of 2 and 4 mg were selected to approximate the systemic exposure of the oral 5 and 10 mg doses (**Supplementary Section S3**). The research pharmacy held the randomization sequence and dispensed visually identical placebo and active preparations, maintaining the blind for participants, raters, and investigators (**Supplementary Section S2**); blinding procedures and integrity are reported in the companion manuscript[14].

### 2.4 Pharmacokinetic Assessments

Plasma Δ9-THC and its metabolites were sampled serially and quantified by liquid chromatography-tandem mass spectrometry; the full pharmacokinetic characterization is reported in the companion manuscript[14].

### 2.5 Quantitative Sensory Testing

Laboratory-evoked pain sensitivity was assessed by a multimodal quantitative sensory testing (QST) battery based on the standardized protocol of the German Research Network on Neuropathic Pain (DFNS)[48]. The QST battery comprised seven threshold and tolerance measures across three stimulus modalities: heat pain threshold (HTH), heat pain tolerance (HTO), cold pain threshold (CTH), and cold pain tolerance (CTO), delivered by contact thermode (TSA-II; Medoc, Ramat Yishai, Israel); pressure pain threshold (PTH) and pressure pain tolerance (PTO), by computer-controlled algometer (AlgoMed; Medoc); and mechanical pain threshold (MTH), by calibrated pinprick stimulators (MRC Systems, Germany). Each measure was obtained twice and averaged. QST was administered at a pre-dose baseline and at five post-dose timepoints (oral arm: 60, 120, 180, 240, and 360 min; vaporized arm: 5, 60, 120, 180, and 360 min). The DFNS protocol governed stimulus application and threshold determination; the tolerance measures and the summary measures defined below were specific to this study. Baseline clinical pain, which was not an eligibility criterion, was characterized at screening with the Brief Pain Inventory-Short Form (BPI-SF)[10].

For each measure, the effect of Δ9-THC was summarized two ways: the session peak change from baseline (the maximum signed deviation across the session) and the session AUC, the area under the change-from-baseline curve integrated by the trapezoidal rule across the post-dose observation window (oral 60-360 min; vaporized 5-360 min). For individual QST measures, session-AUC estimates and confidence intervals were divided by the window duration and are reported as mean changes from baseline in each measure’s original units. For the cold measures (CTH, CTO), the change score was sign-inverted so that positive values denote analgesia (greater cold tolerated) across every measure.

### 2.6 Pain Sensitivity Index

As a data reduction strategy, we combined the four thermal QST measures (HTH, HTO, CTH, CTO) into a single pain sensitivity index. This approach is not only consistent with previous integration of QST measures into composite indices[7,26], but it also limits multiplicity across correlated measures and averages out measurement noise. The combined measures all share a transduction substrate: noxious heat and cold are detected by TRP channels on Aδ and C nociceptors[28]. CB1 receptors are co-expressed with TRPV1 on those same afferents, where a locally applied CB1 agonist attenuates thermal hyperalgesia[5]. We built the pain sensitivity index on session AUCs rather than peak changes because, in laboratory-evoked pain models, analgesic effects are detected more robustly by the area under the pain curve than by peak pain intensity[51]. For each session, the four session AUCs were sign-inverted for the cold measures so that positive values denote analgesia, standardized within measure as z-scores across all sessions of both arms. The pain sensitivity index is therefore in SD units, one unit being an average change of one standard deviation across its four component measures. All QST measures are also reported separately.

### 2.7 Abuse Liability

Subjective drug effects were the primary measure of abuse liability and behavioral reinforcement value served as a secondary measure. To assess subjective drug effects, we used the Pleasurable composite of the Drug Effects Questionnaire (DEQ), a visual analog scale with established psychometric properties[41]. The Pleasurable composite was the mean of three items (like the drug effects, feel good effects, want more drug), which map onto measures named in the FDA guidance Assessment of Abuse Potential of Drugs[52], and was summarized as the peak change from baseline (E_max_), the summary that guidance recommends for primary analyses of abuse potential, and as AUC_0-8h_, parallel to the QST outcomes.

To assess reinforcement value, we administered the Multiple-Choice Procedure once per session, at 480 min after dosing. Participants made a series of choices between a repeat of that session’s dose and increasing amounts of money, and the outcome was the crossover point: the lowest amount of money chosen over the assigned dose for that test session[20].

### 2.8 Statistical Analysis

QST outcomes were summarized as the area under the change-from-baseline curve across the session and analyzed using linear mixed models. The oral and vaporized arms were analyzed separately. Dose and period (first, second or third session) were fixed effects, the latter to account for practice effects, and dose was a within-participant repeated factor under an unstructured variance-covariance matrix. Denominator degrees of freedom used the Kenward-Roger approximation[29]. The pain sensitivity index was analyzed identically. For peak DEQ Pleasurable and the Multiple-Choice Procedure crossover point, we report the companion manuscript’s estimates, from the same model under a heterogeneous compound-symmetry variance-covariance matrix. Pairwise dose contrasts are reported as least-squares mean differences with 95% confidence intervals in original units, accompanied by Hedges’ g computed from the model-estimated standard deviations (SDs) of the two doses compared[31]. A within-arm low-versus-high contrast tested the biphasic-dose hypothesis. Given the exploratory, proof-of-concept design, pairwise contrasts are reported for every outcome regardless of the omnibus test, and p-values were not corrected for multiple comparisons[49]. Analyses were performed in R version 4.5.2 with two-sided α = 0.05.

Additionally, we asked whether analgesia and abuse liability track drug exposure rather than administered dose. We related both outcomes to each session’s peak concentration (C_max_) and total exposure (AUC_0-8h_) of Δ9-THC plus 11-OH-THC[12,57], using linear mixed models with a random intercept for participant, which accounts for the correlation among each participant’s sessions (**Supplementary Section S8).**

Finally, we compared the conditions of both routes on their model-estimated analgesia and abuse liability using a benefit-risk Pareto frontier, an approach that has guided dose selection in tuberculosis[37] and cancer combination therapy[18]. We refer to our application as the Analgesia-Abuse Liability Frontier (AALF; **Supplementary Section S5**). Time courses are shown descriptively and were not tested statistically (**Supplementary Section S9**).

## 3. Results

### 3.1 Participants

Twenty older adults were enrolled, 10 per arm (**Table 1**). Overall, mean age was 70.0 (SD: 5.1) years, 30% (n=6) were female, and mean body mass index (BMI) was 28.6 (SD: 4.2) kg/m². Baseline clinical pain was low on the BPI-SF: mean severity 1.5 (SD: 1.6) and interference 1.1 (SD: 2.0). The two arms were balanced on demographic, baseline pain, and cannabis use characteristics (**Table 1**, all p≥.20). In the oral arm, 20% (n=2) were female, mean age was 70.5 (SD: 4.7) years, and mean BMI was 27.6 (SD: 4.2) kg/m². In the vaporized arm, 40% (n=4) were female, mean age was 69.5 (SD: 5.7) years, and mean BMI was 29.6 (SD: 4.2) kg/m². One vaporized-arm participant was withdrawn after a vasovagal episode during the first test session.

**Table 1.** Participant characteristics by treatment arm.

| **Characteristic** |  | **Treatment arm** |  |  |
|---|---|---|---|---|
|  | **Overall**<br>**N = 20** | **Oral**<br>**N = 10** | **Vaporized**<br>**N = 10** | **p-value** |
| **Age, years** | 70.0 (5.1) | 70.5 (4.7) | 69.5 (5.7) | .67 |
| **Sex (male)** | 14 (70%) | 8 (80%) | 6 (60%) | .63 |
| **Race** |  |  |  | >.99 |
| Black | 0 (0%) | 0 (0%) | 0 (0%) |  |
| Asian | 0 (0%) | 0 (0%) | 0 (0%) |  |
| Native American | 0 (0%) | 0 (0%) | 0 (0%) |  |
| White | 19 (95%) | 10 (100%) | 9 (90%) |  |
| Other | 1 (5.0%) | 0 (0%) | 1 (10%) |  |
| **Ethnicity** |  |  |  | >.99 |
| Hispanic | 1 (5.0%) | 0 (0%) | 1 (10%) |  |
| Not Hispanic | 19 (95%) | 10 (100%) | 9 (90%) |  |
| **Height, cm** | 171.6 (12.1) | 170.9 (16.3) | 172.2 (6.5) | .82 |
| **Weight, kg** | 84.5 (16.7) | 80.7 (16.5) | 88.2 (16.9) | .33 |
| **Body mass index, kg/m<sup>2</sup>** | 28.6 (4.2) | 27.6 (4.2) | 29.6 (4.2) | .31 |
| **Body fat, %** | 30.5 (4.4) | 28.8 (2.7) | 31.8 (5.1) | .22 |
| **BPI-SF pain severity** | 1.5 (1.6) | 1.8 (2.2) | 1.2 (0.9) | .49 |
| **BPI-SF pain interference** | 1.1 (2.0) | 1.5 (2.8) | 0.8 (0.8) | .45 |
| **Age at first cannabis use, years** | 17.0 (2.8) | 17.2 (2.9) | 16.7 (2.9) | .70 |
| **Total years of cannabis use** | 33.6 (19.2) | 33.6 (19.7) | 33.7 (19.9) | .99 |
| **Past 30-day use (0-8 ordinal)** | 3.4 (2.9) | 3.4 (2.7) | 3.3 (3.2) | .94 |
| **Lifetime days of cannabis use** |  |  |  | >.99 |
| 0 | 0 (0%) | 0 (0%) | 0 (0%) |  |
| 1-10 | 3 (15%) | 1 (10%) | 2 (20%) |  |
| 11-50 | 0 (0%) | 0 (0%) | 0 (0%) |  |
| 51-100 | 1 (5.0%) | 1 (10%) | 0 (0%) |  |
| 101-200 | 1 (5.0%) | 0 (0%) | 1 (10%) |  |
| 201-300 | 1 (5.0%) | 1 (10%) | 0 (0%) |  |
| >300 | 14 (70%) | 7 (70%) | 7 (70%) |  |
| **Typical method of consumption** |  |  |  | .20 |
| Joint | 9 (47%) | 7 (70%) | 2 (22%) |  |
| Bowl | 2 (11%) | 1 (10%) | 1 (11%) |  |
| Bong | 2 (11%) | 1 (10%) | 1 (11%) |  |
| One-hitter | 2 (11%) | 0 (0%) | 2 (22%) |  |
| Ingestion (e.g. food) | 4 (21%) | 1 (10%) | 3 (33%) |  |

Mean (SD) for continuous variables; n (%) for categorical variables. Between-arm tests are Welch’s two-sample t-test for continuous variables and Fisher’s exact test for categorical variables, without multiplicity correction (balance check, exploratory). Body fat percentage was available for 12 of 20 person-arms (oral, 5 of 10; vaporized, 7 of 10) and is summarized on available data. BPI-SF, Brief Pain Inventory-Short Form.

### 3.2 Pain Thresholds and Tolerance

We observed a dose effect of Δ9-THC on four of the seven QST measures. In the oral arm, dose significantly affected heat pain threshold (p=.025) and pressure pain threshold (p=.020; **Table 2**, **Figure 1**). The 5 mg dose raised heat pain threshold above both placebo (g=0.75, p=.041) and 10 mg (g=0.57, p=.046), and pressure pain threshold above 10 mg (g=0.86, p=.018). In the vaporized arm, dose significantly affected heat pain tolerance (p=.017) and mechanical pain threshold (p=.011). The 2 mg dose raised heat pain tolerance above 4 mg (g=0.55, p=.006) and mechanical pain threshold above placebo (g=1.25, p=.010). Across both arms, all five significant pairwise contrasts favored the lower doses, and neither higher dose differed significantly from placebo on any measure.

**Figure 1.**
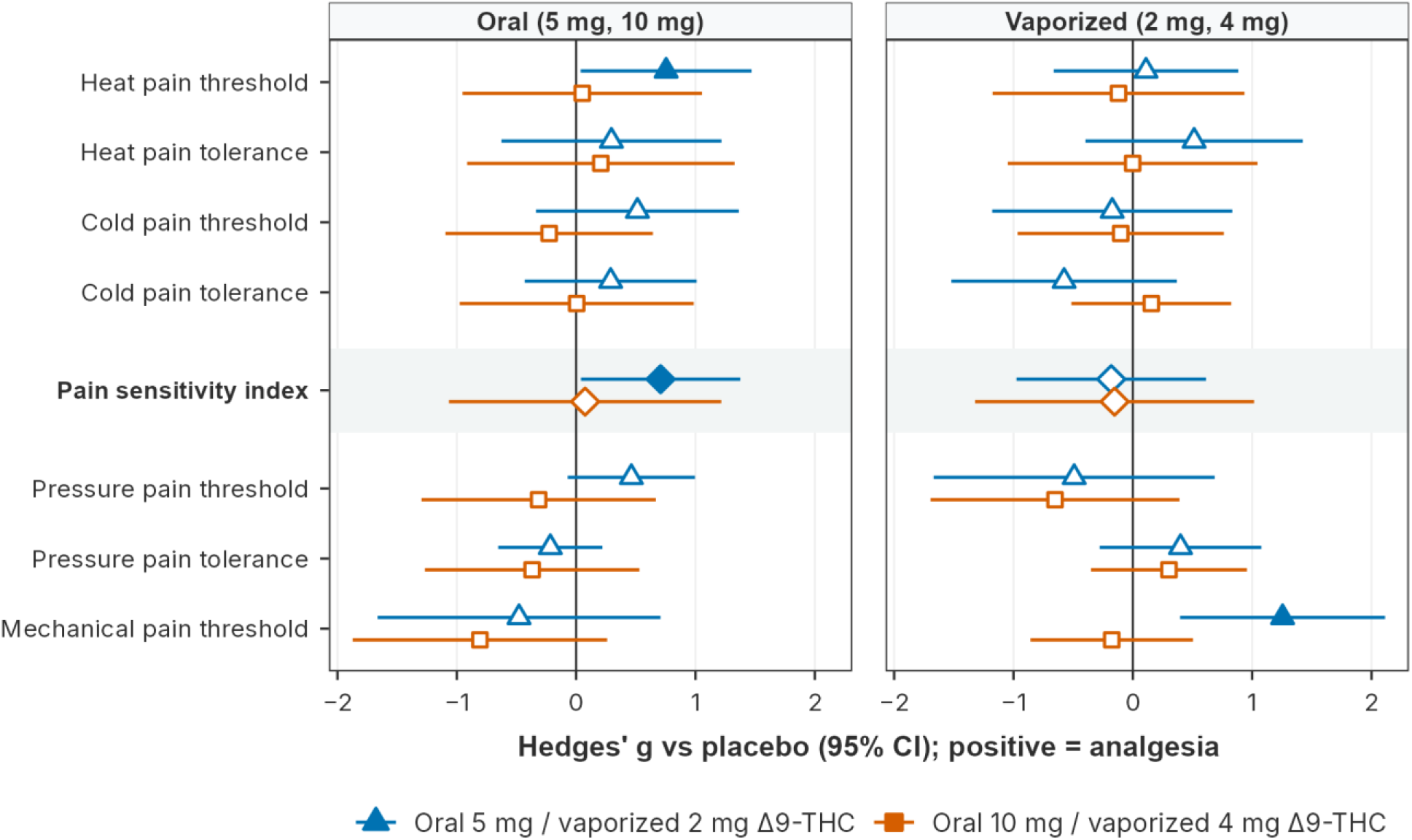
Effects of Δ9-THC on laboratory-evoked pain, by measure and route. Points are Hedges’ g for each dose against placebo, and bars are the 95% confidence intervals of the model-estimated mean differences, expressed in g units (**Table 2**). Blue triangles, lower dose; orange squares, higher dose. Positive values denote analgesia; cold thresholds and tolerances are sign-inverted. Filled symbols mark contrasts significant at p < .05. The four thermal measures (top) are combined into the pain sensitivity index (diamonds, shaded row); the pressure and mechanical measures follow.

**Table 2.**
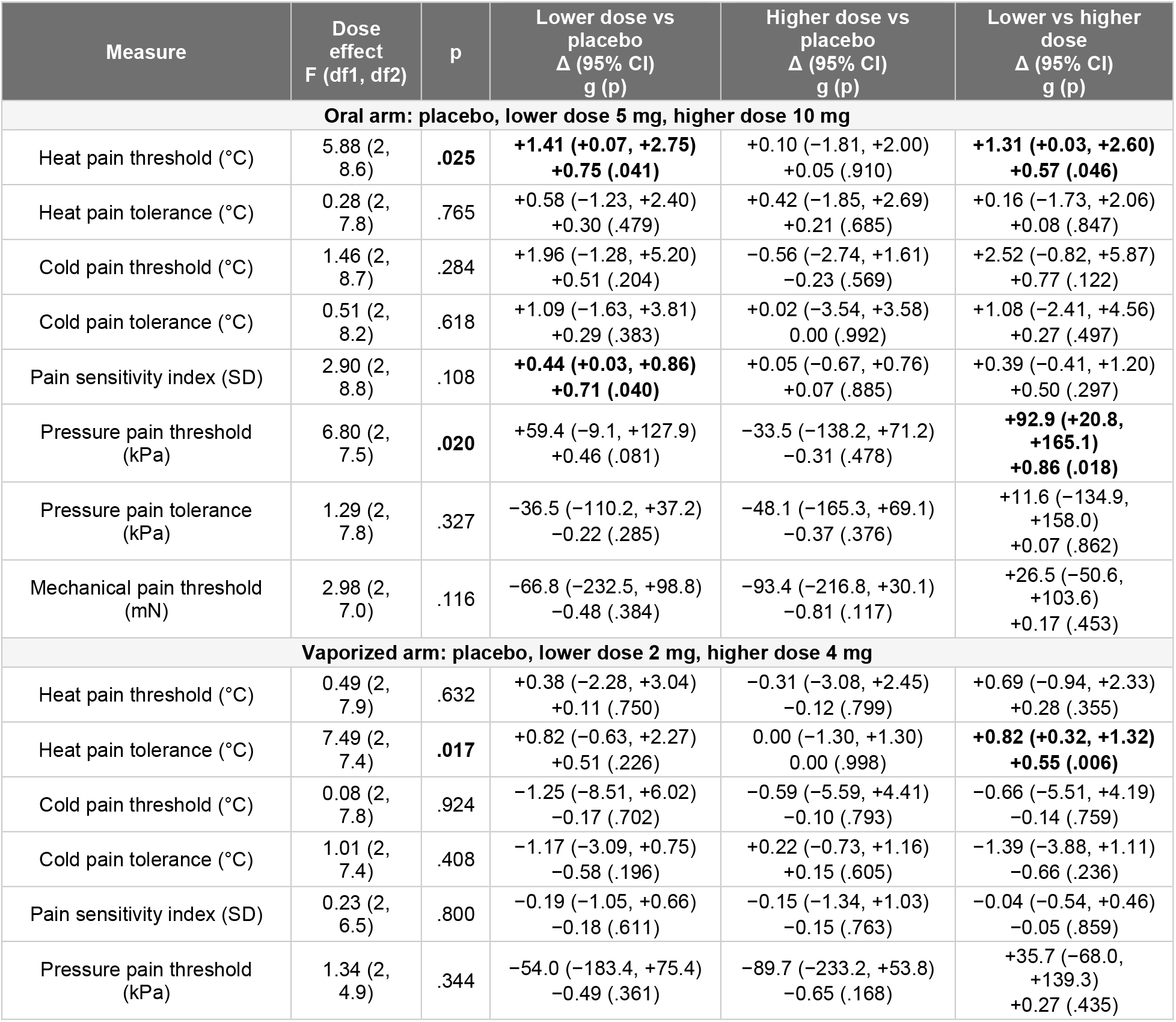
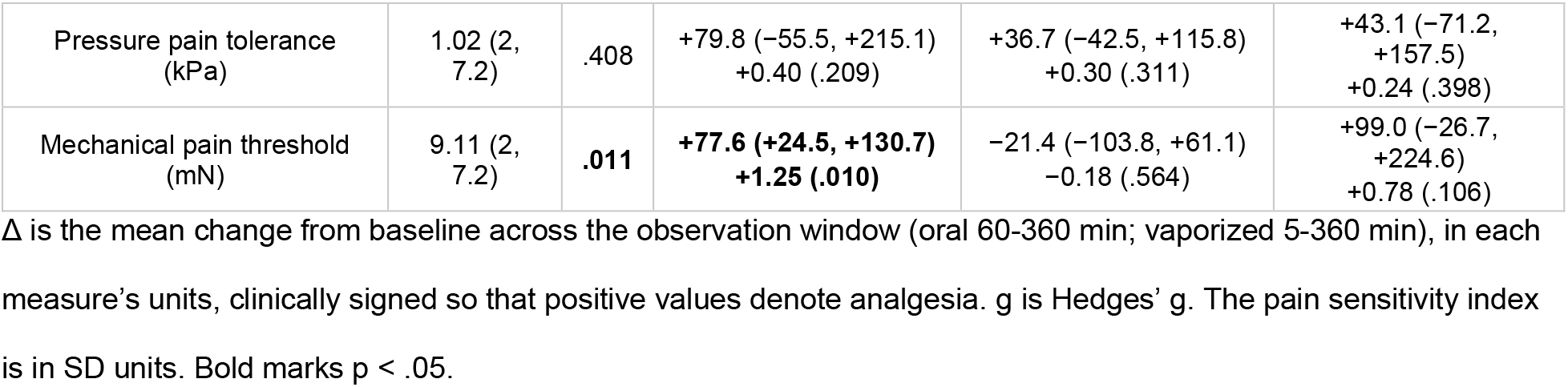
Dose effects on quantitative sensory testing, by arm and measure.

| Measure | Dose effect<br>F (df1, df2) | p | Lower dose vs placebo<br>Δ (95% CI)<br>g (p) | Higher dose vs placebo<br>Δ (95% CI)<br>g (p) | Lower vs higher dose<br>Δ (95% CI)<br>g (p) |
|---|---|---|---|---|---|
| Oral arm: placebo, lower dose 5 mg, higher dose 10 mg |  |  |  |  |  |
| Heat pain threshold (°C) | 5.88 (2, 8.6) | **.025** | **+1.41 (+0.07, +2.75)<br>+0.75 (.041)** | +0.10 (−1.81, +2.00)<br>+0.05 (.910) | **+1.31 (+0.03, +2.60)<br>+0.57 (.046)** |
| Heat pain tolerance (°C) | 0.28 (2, 7.8) | .765 | +0.58 (−1.23, +2.40)<br>+0.30 (.479) | +0.42 (−1.85, +2.69)<br>+0.21 (.685) | +0.16 (−1.73, +2.06)<br>+0.08 (.847) |
| Cold pain threshold (°C) | 1.46 (2, 8.7) | .284 | +1.96 (−1.28, +5.20)<br>+0.51 (.204) | −0.56 (−2.74, +1.61)<br>−0.23 (.569) | +2.52 (−0.82, +5.87)<br>+0.77 (.122) |
| Cold pain tolerance (°C) | 0.51 (2, 8.2) | .618 | +1.09 (−1.63, +3.81)<br>+0.29 (.383) | +0.02 (−3.54, +3.58)<br>0.00 (.992) | +1.08 (−2.41, +4.56)<br>+0.27 (.497) |
| Pain sensitivity index (SD) | 2.90 (2, 8.8) | .108 | **+0.44 (+0.03, +0.86)<br>+0.71 (.040)** | +0.05 (−0.67, +0.76)<br>+0.07 (.885) | +0.39 (−0.41, +1.20)<br>+0.50 (.297) |
| Pressure pain threshold (kPa) | 6.80 (2, 7.5) | **.020** | +59.4 (−9.1, +127.9)<br>+0.46 (.081) | −33.5 (−138.2, +71.2)<br>−0.31 (.478) | **+92.9 (+20.8, +165.1)<br>+0.86 (.018)** |
| Pressure pain tolerance (kPa) | 1.29 (2, 7.8) | .327 | −36.5 (−110.2, +37.2)<br>−0.22 (.285) | −48.1 (−165.3, +69.1)<br>−0.37 (.376) | +11.6 (−134.9, +158.0)<br>+0.07 (.862) |
| Mechanical pain threshold (mN) | 2.98 (2, 7.0) | .116 | −66.8 (−232.5, +98.8)<br>−0.48 (.384) | −93.4 (−216.8, +30.1)<br>−0.81 (.117) | +26.5 (−50.6, +103.6)<br>+0.17 (.453) |
| Vaporized arm: placebo, lower dose 2 mg, higher dose 4 mg |  |  |  |  |  |
| Heat pain threshold (°C) | 0.49 (2, 7.9) | .632 | +0.38 (−2.28, +3.04)<br>+0.11 (.750) | −0.31 (−3.08, +2.45)<br>−0.12 (.799) | +0.69 (−0.94, +2.33)<br>+0.28 (.355) |
| Heat pain tolerance (°C) | 7.49 (2, 7.4) | **.017** | +0.82 (−0.63, +2.27)<br>+0.51 (.226) | 0.00 (−1.30, +1.30)<br>0.00 (.998) | **+0.82 (+0.32, +1.32)<br>+0.55 (.006)** |
| Cold pain threshold (°C) | 0.08 (2, 7.8) | .924 | −1.25 (−8.51, +6.02)<br>−0.17 (.702) | −0.59 (−5.59, +4.41)<br>−0.10 (.793) | −0.66 (−5.51, +4.19)<br>−0.14 (.759) |
| Cold pain tolerance (°C) | 1.01 (2, 7.4) | .408 | −1.17 (−3.09, +0.75)<br>−0.58 (.196) | +0.22 (−0.73, +1.16)<br>+0.15 (.605) | −1.39 (−3.88, +1.11)<br>−0.66 (.236) |
| Pain sensitivity index (SD) | 0.23 (2, 6.5) | .800 | −0.19 (−1.05, +0.66)<br>−0.18 (.611) | −0.15 (−1.34, +1.03)<br>−0.15 (.763) | −0.04 (−0.54, +0.46)<br>−0.05 (.859) |
| Pressure pain threshold (kPa) | 1.34 (2, 4.9) | .344 | −54.0 (−183.4, +75.4)<br>−0.49 (.361) | −89.7 (−233.2, +53.8)<br>−0.65 (.168) | +35.7 (−68.0, +139.3)<br>+0.27 (.435) |
| Pressure pain tolerance (kPa) | 1.02 (2, 7.2) | .408 | +79.8 (−55.5, +215.1)<br>+0.40 (.209) | +36.7 (−42.5, +115.8)<br>+0.30 (.311) | +43.1 (−71.2, +157.5)<br>+0.24 (.398) |
| Mechanical pain threshold (mN) | 9.11 (2, 7.2) | **.011** | **+77.6 (+24.5, +130.7)<br>+1.25 (.010)** | −21.4 (−103.8, +61.1)<br>−0.18 (.564) | +99.0 (−26.7, +224.6)<br>+0.78 (.106) |

Δ is the mean change from baseline across the observation window (oral 60-360 min; vaporized 5-360 min), in each measure’s units, clinically signed so that positive values denote analgesia. g is Hedges’ g. The pain sensitivity index is in SD units. Bold marks p < .05.

### 3.3 Pain Sensitivity Index

We observed no significant overall dose effect on the pain sensitivity index in either arm (oral p=.108; vaporized p=.800; **Figure 2**). Within the oral arm, however, the pairwise contrast of 5 mg against placebo was significant (mean difference: 0.44 SD, 95% CI 0.03 to 0.86; g=0.71, p=.040), whereas 10 mg did not differ from placebo (0.05 SD; g=0.07, p=.885). Neither vaporized dose differed from placebo (2 mg: g=−0.18, p=.611; 4 mg: g=−0.15, p=.763).

**Figure 2.**
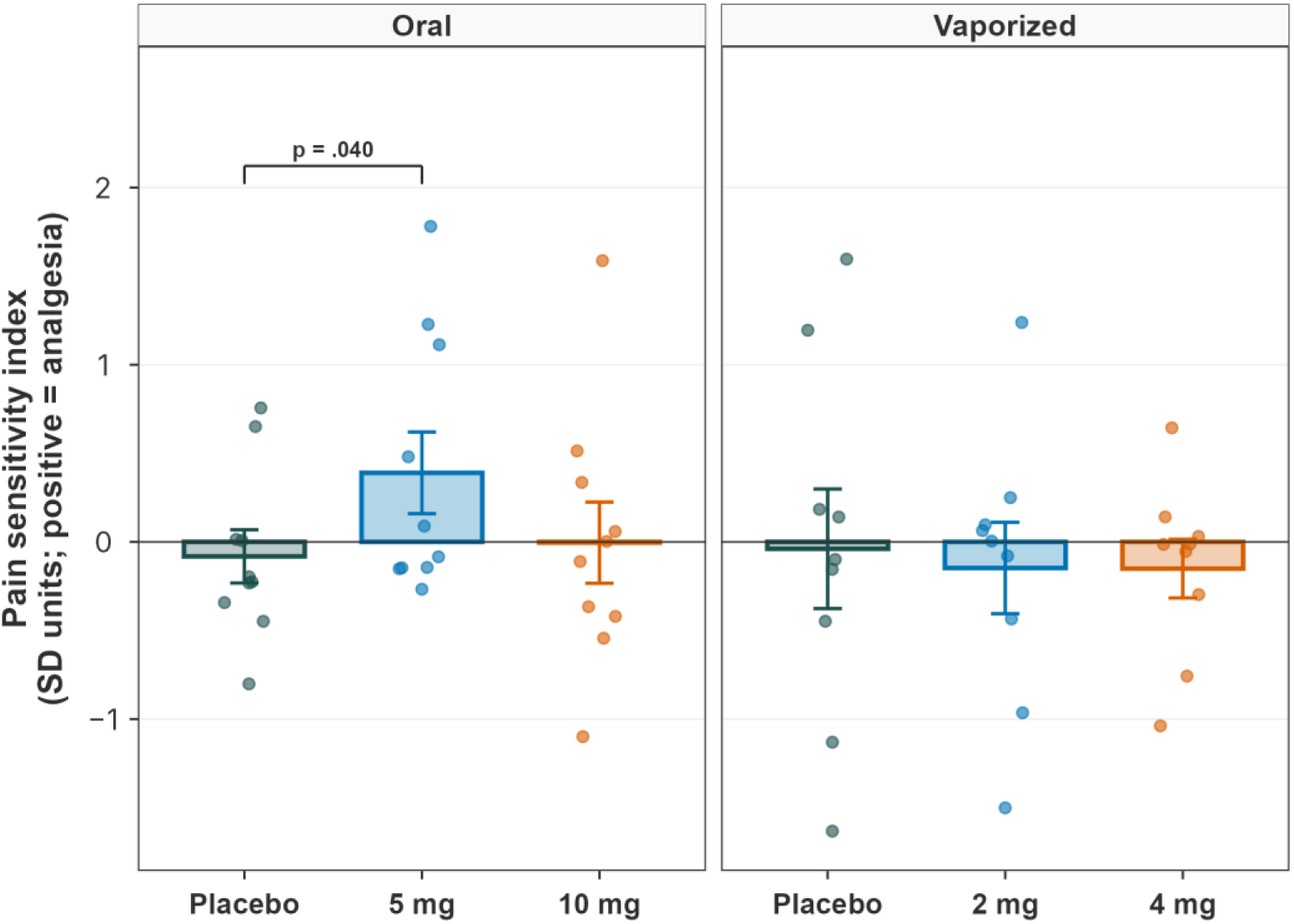
Pain sensitivity index by dose and route. Values are summarized as the area under the change-from-baseline curve across the session and expressed in SD units. Bars, group means ± SE; points, individual sessions. Brackets mark contrasts significant at p < .05.

### 3.4 Abuse Liability

The dose effect on peak DEQ Pleasurable was significant in both arms (oral p=.038; vaporized p=.019; **Supplementary Figure S2**). In the oral arm, both doses raised it above placebo (5 mg: +19.1 points, 95% CI 3.4 to 34.8; g=0.84, p=.021; 10 mg: +22.8, 95% CI 2.0 to 43.6; g=0.77, p=.034). In the vaporized arm, 2 mg did not differ significantly from placebo (+21.0, 95% CI −9.1 to 51.0; g=0.68, p=.153), whereas 4 mg did (+35.1, 95% CI 12.2 to 58.1; g=1.49, p=.007). DEQ Pleasurable rose faster and higher after vaporization than after oral administration (**Figure 3**). Reinforcement value showed the same route asymmetry. In the oral arm, neither the dose effect (p=.670) nor contrast against placebo (5 mg: −$0.4, p=.911; 10 mg: +$2.1, p=.533) was significant. In the vaporized arm, the dose effect was significant (p=.018): 2 mg did not differ significantly from placebo (+$4.8, 95% CI −0.1 to 9.7; g=0.70, p=.052), whereas 4 mg raised the crossover point above placebo by $9.0 (95% CI 3.6 to 14.5; g=1.05, p=.004). When both outcomes were considered together, no other condition produced more analgesia than oral 5 mg at no greater abuse liability (**Supplementary Section S5**).

**Figure 3.**
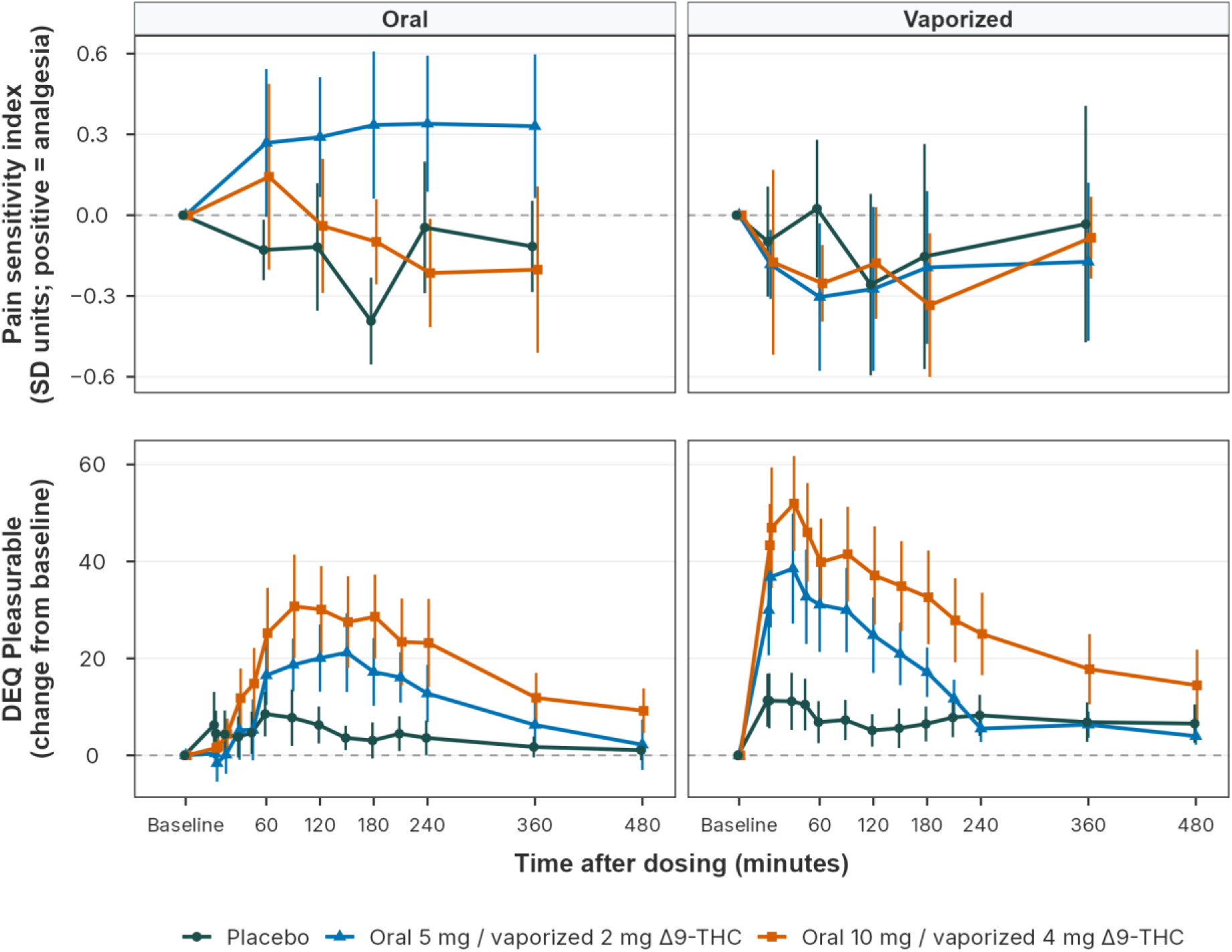
Pain sensitivity index and subjective drug effects over time, by route and dose. Top, the pain sensitivity index at each assessment, on the same SD scale as Figure 2; positive values denote analgesia. Bottom, change from baseline in the Drug Effects Questionnaire Pleasurable composite. Points, group means; bars, standard errors. The pain sensitivity index is shown through 360 min, the end of the window over which it is computed.

### 3.5 Time Course of Analgesia and Subjective Effects

After oral 5 mg, the pain sensitivity index showed more analgesia than placebo at every assessment from 60 to 360 min (by 0.39 to 0.73 SD; **Figure 3**). DEQ Pleasurable peaked at 150 min and was 4.5 points above placebo at 360 min. After oral 10 mg, the pain sensitivity index showed more analgesia than placebo through 180 min (by 0.09 to 0.29 SD) and less thereafter (by 0.17 and 0.09 SD). DEQ Pleasurable peaked at 90 min and was 10.1 points above placebo at 360 min.

After vaporized 2 mg, the pain sensitivity index showed less analgesia than placebo at every assessment (by 0.02 to 0.33 SD). DEQ Pleasurable peaked at 30 min and was 0.5 points below placebo at 360 min. After vaporized 4 mg, the pain sensitivity index differed from placebo by −0.28 to +0.08 SD. DEQ Pleasurable peaked at 30 min and was 10.9 points above placebo at 360 min.

### 3.6 Relationships Between Exposure, Analgesia, and Abuse Liability

Both abuse liability measures increased with combined Δ9-THC and 11-OH-THC exposure. DEQ Pleasurable increased by 14.5 points per SD of C_max_ (p<.001) and by 10.6 points per SD of AUC_0-8h_ (p=.003). Reinforcement value increased by $1.97 per SD of C_max_ (p=.045) and by $1.98 per SD of AUC_0-8h_ (p=.044). No significant association between analgesia and either exposure metric was detected (p=.585 and p=.903; **Supplementary Section S8**).

### 3.7 General Safety and Adverse Events

Adverse events occurred only at vaporized 4 mg. Five events were recorded in two participants, including a severe vasovagal episode that prompted withdrawal. No adverse events were recorded across the 30 oral sessions. Detailed safety, cardiovascular, and tolerability outcomes are reported in the companion manuscript[14].

## 4. Discussion

In this randomized, double-blind, placebo-controlled trial in adults aged 65 and older, analgesic effects were detected on selected QST measures at the lower dose of each route. Oral 5 mg increased heat pain threshold and the pain sensitivity index relative to placebo, whereas vaporized 2 mg increased mechanical pain threshold. Neither higher dose, oral or vaporized, differed significantly from placebo on any QST measure. Subjective pleasurable drug effects increased at both oral doses and at vaporized 4 mg, whereas reinforcement value increased significantly only after vaporized 4 mg. Oral 5 mg showed the most favorable observed balance between analgesia and abuse liability. It was the only condition whose pain sensitivity index showed more analgesia than placebo at every assessment, and its peak pleasurable response was the smallest of any active condition. It was also the only active condition on the benefit-risk Pareto frontier, which needs no weighting of one outcome against the other (**Supplementary Section S5**). This result highlights the relative advantage of the oral 5 mg compared to the other tested dose-route conditions, but should not be viewed as evidence of an optimal dose. All recorded adverse events occurred after vaporized 4 mg.

### 4.1 Implications for Understanding the Analgesic Effects and Abuse Liability of Δ9-THC Among Older Adults

The analgesic response pattern was suggestive of an inverted U, rather than a monotonic increase with dose. Every significant low-versus-high contrast favored the lower dose: oral 5 mg over 10 mg on heat and pressure pain thresholds, and vaporized 2 mg over 4 mg on heat pain tolerance. Biphasic dose-response has been documented for Δ9-THC’s anxiety-related effects: in healthy young adults, 7.5 mg of oral Δ9-THC reduced subjective distress after a psychosocial stressor, whereas 12.5 mg increased anxiety and negative mood[9]. For analgesia, the evidence is sparser but suggests a similar relationship. In healthy volunteers, smoked cannabis containing 4% Δ9-THC reduced capsaicin-induced pain, whereas 8% increased it and 2% had no effect[53]. A single 20 mg oral dose of Δ9-THC likewise produced hyperalgesia rather than analgesia on heat and cold testing[43].

The abuse liability of Δ9-THC among older adults is contingent not only on dose, but also on route of administration. Vaporization produced the faster and larger subjective response in this trial and, at 4 mg, the only increase in reinforcement value. In a laboratory study of adults with a mean age of 68.7 years, smoked cannabis likewise kept subjective ratings elevated for five hours[16]. A review of cannabis and aging found few and inconsistent studies in older adults[56], and no prior controlled study in adults aged 65 and older had assessed analgesia and abuse liability together across doses and routes. Taken together, these findings demonstrate measurable abuse liability of Δ9-THC and cannabis in older adults, and support jointly assessing analgesia and abuse liability of cannabinoids in this population.

### 4.2 Comparing Doses and Routes on Analgesia and Abuse Liability

After oral Δ9-THC, the duration of analgesia extended beyond the peak of abuse-related effects. Pleasurable effects after oral 5 mg peaked at 150 min and had nearly returned to placebo levels by 360 min. The pain sensitivity index, by contrast, showed more analgesia than placebo from 60 through 360 min, and the difference was largest at 180 min. An earlier study that compared oral and smoked Δ9-THC on both outcomes in 30 adults who used cannabis daily (mean age 27 years) found the same pattern[13]. In that study, abuse-related ratings after oral dronabinol peaked at 60 to 90 min, whereas analgesia peaked at 180 min. Peak analgesia on the cold pressor test was similar for oral dronabinol and smoked cannabis, but dronabinol’s analgesia lasted longer and its abuse-related ratings were lower[13].

Our results differed from those of the earlier study on dose. Both outcomes changed at a lower oral dose in our study: 5 mg raised pleasurable effects and produced analgesia. In the earlier study, dronabinol raised abuse-related ratings and reduced pain sensitivity only at 20 mg[13]. Tolerance from daily cannabis use[11] and age-related changes in the pharmacokinetics and pharmacodynamics of Δ9-THC[15] may both contribute. The pain tests also differed. The earlier study measured pain while participants held one hand in 4 °C water (the cold pressor test). Our pain sensitivity index combined heat and cold pain thresholds and tolerances measured with a contact thermode.

### 4.3 Implications for Understanding the Relationship Between Exposure to Δ9-THC, Analgesia, and Abuse Liability Outcomes

Both abuse liability measures tracked peak and total exposure to Δ9-THC and its active metabolite, whereas no significant association between analgesia and either exposure metric was detected. A significant association for abuse liability and a null for analgesia do not establish that the two outcomes depend on different features of the concentration-time profile: the analgesic estimates were imprecise, and the study was not designed to separate peak (C_max_) from total (AUC_0-8h_) exposure, which co-vary within each route and participant. The data show only that the active condition with the lowest C_max_, oral 5 mg, was associated with the most analgesia, while abuse liability rose with exposure. Whether analgesic effects are relatively insensitive to exposure differences within this range may be evaluated in a trial designed to dissociate peak from total exposure, for example by varying the rate of delivery at a fixed dose.

### 4.4 Limitations

Several limitations bound our results. First, the sample was small, predominantly male and White, and composed entirely of healthy volunteers with prior cannabis exposure, although all tested negative for recent use at enrollment; generalizability to cannabis-naive older adults is therefore limited. Second, the pain model: evoked pain thresholds may not always translate into relief of chronic pain[45], although a systematic review found broadly similar cannabinoid response patterns in healthy volunteers and patients with chronic non-cancer pain[42]. Third, the design: between-participant variability may confound cross-route comparisons, and single-dose sessions cannot establish sustained efficacy or characterize tolerance[11]. In the earlier study, smoked cannabis produced analgesia that peaked at 15 min[13]. The QST schedule here had no assessment between 5 and 60 min after vaporization, so it could have missed an effect this brief. Finally, the analysis: with seven QST measures and multiple contrasts, some significant results may be chance findings, because analyses were exploratory and uncorrected for multiplicity; the consistent direction of effects across measures, rather than any single p-value, is the finding to weigh.

### 4.5 Future Directions

These findings support evaluating oral 5 mg Δ9-THC in repeated-dose trials involving older adults with chronic pain. Such trials should determine whether the observed laboratory analgesic effects translate into sustained improvements in clinical pain and function, while assessing how abuse-related responses change with continued treatment and how they differ between cannabis-naive and cannabis-experienced older adults. Given the association between chronic pain and CUD[25], such trials should also measure the emergence of problematic use alongside analgesia and tolerability. Additional doses below and around 5 mg would help determine whether the favorable observed balance extends across a dose range. Studies that distinguish the contributions of peak and total exposure could also guide delivery strategies. If analgesia is preserved while abuse liability is reduced by lowering peak concentration independently of total exposure, a delivery profile that maintains total exposure while limiting peaks could improve the benefit-risk balance. Repeated low doses of inhaled Δ9-THC could, in principle, approximate the profile of oral 5 mg where rapid onset matters, as in breakthrough pain[4]; whether they would preserve analgesia is unknown. The benefit-risk frontier could also be applied to other cannabinoids and to balanced Δ9-THC:CBD formulations.

## 5. Conclusion

In this randomized, double-blind, placebo-controlled trial in adults aged 65 and older, we compared two doses each of oral and vaporized Δ9-THC on laboratory-evoked pain and abuse liability, measured together across each session. Significant analgesic effects were confined to the lower dose of each route, consistent with the hypothesized inverted-U dose-response; oral 5 mg offered the most analgesia for the least abuse liability; and adverse events occurred only at vaporized 4 mg. These exploratory findings make oral 5 mg the priority for trials in older adults with chronic pain that assess analgesia, abuse liability, and tolerability together.

## Data Availability

De-identified data and the R code used for all analyses are available from the corresponding author upon reasonable request, under a data use agreement.

## Acknowledgments

We thank Christina Riggione, Jocelyn Ra, Enoch Ofori, Mayte A. Cerezo-Matias, and Becky Suh for their contributions to data collection. We are grateful to Susan Gray, BSN, RN, Angelina Genovese, RNC, BSN, MBA, and Vanessa Hurley, RN, for nursing support. We also thank the participants and the staff of the Biological Studies Unit at the VA Connecticut Healthcare System.

## Use of generative AI

Claude (Anthropic) was used to assist with the data analysis code and to proofread parts of the manuscript. The authors reviewed all output and take full responsibility for the content.

