## Supplementary Material for "Balancing Analgesic Effects and Abuse Liability in Older Adults: A Randomized, Placebo-Controlled Trial of Oral and Vaporized Δ9-Tetrahydrocannabinol"

### S1. Inclusion and Exclusion Criteria

**Inclusion criteria:**

1. Healthy adults aged 65 years or older.
2. Prior exposure to Δ9-THC or cannabis: at least once in the last 10 years; 1-10 times in the last 20 years; or more than 20 times lifetime.
3. Capable of providing written informed consent in English.

**Exclusion criteria:**

1. Meeting DSM-5 criteria for a psychiatric or substance use disorder, other than tobacco use disorder, within the last year.
2. Current cannabinoid use, evidenced by urine drug screen.
3. History of treatment for cannabis use disorder, or history of intent or current intent of abstaining from cannabis use.
4. Clinically significant medical disorders, including liver or kidney dysfunction, chronic obstructive pulmonary disease or asthma, immunosuppressing conditions, and history or presence of epilepsy, seizures, or head trauma with loss of consciousness.
5. Neurological conditions that may change the response to nociceptive stimuli, such as stroke or peripheral neuropathy, or that lead to loss of balance, evidenced by a neuro-sensory examination.
6. Contraindications for exposure to nociceptive stimuli, such as untreated hypertension.
7. Current regular use of drugs known to affect pain, or that are prominent inducers or inhibitors of CYP2C9, CYP3A4, or UGT1A9, such as carbamazepine, valproate, fluvoxamine, and paroxetine.
8. Major neurocognitive disorders precluding participation, evidenced by clinical examination.
9. Abnormal electrocardiogram, arrhythmia, vasospastic disease, chronic heart failure, or presence of a pacemaker.
10. Elevation of liver enzymes (ALT or AST) to twice the upper limit of normal or higher.
11. Personal or family history of a primary psychotic disorder, or a mood disorder with psychotic features.
12. Current suicidal ideation.
13. Allergy or serious adverse reaction to sesame oil, Δ9-THC, or cannabis.
14. Receipt of any drug as part of a research study within 30 days before study medication administration.
15. Medical conditions that increase the risk of respiratory problems, such as chronic obstructive pulmonary disease, asthma, recurring bronchitis, or reactive airway disorder (vaporized arm only).
16. History of environmental sensitivities, such as bronchospastic allergies or multiple chemical sensitivities, or other airway sensitivities that require an epinephrine auto-injector (vaporized arm only).

### S2. Blinding and Allocation Concealment

Conditions were delivered in counterbalanced random order across three test days per arm, at least 72 hours apart. The research pharmacy held the randomization sequence and dispensed visually identical placebo and active preparations (over-encapsulated capsules; a matched ethanolic vehicle delivered through the same vaporizer and paced-puffing procedure). Participants, raters, and investigators were blinded on each test day. Preparation procedures and the full blinding assessment are reported in the companion manuscript.

#### S3. Exposure Overlap Between Routes

**Figure S1** shows peak concentration and total exposure of the active combined analytes (Δ9-THC plus 11-OH-THC) for every active session. The routes overlapped on total exposure, each vaporized median falling within the range of both oral doses, but separated on peak concentration, where the vaporized doses ran roughly four- to five-fold higher than the matched oral doses (nine-fold on Δ9-THC alone, companion manuscript).

**Figure S1. Peak concentration ($C_{max}$, left) and total exposure ($AUC_{0\text{-}8h}$, right) of the active combined analytes (Δ9-THC + 11-OH-THC) by dose and route.** Boxes, median and interquartile range; points, individual sessions; logarithmic x-axis.

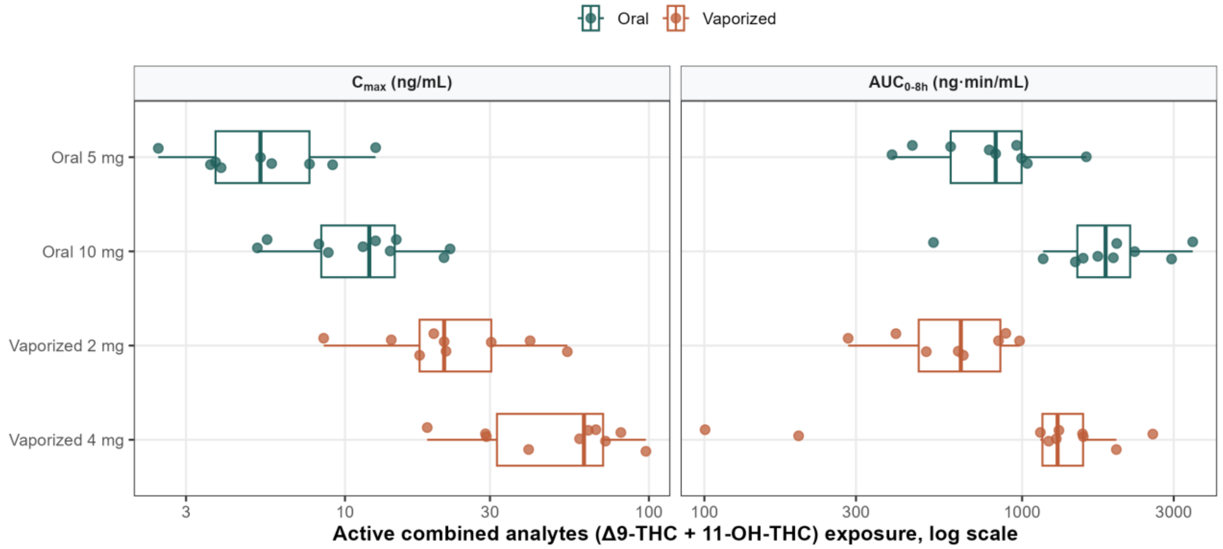

**S4. Peak Subjective Effects**

**Figure S2** shows the peak change from baseline in DEQ Pleasurable ($E_{max}$), the summary behind the abuse liability contrasts in main text Section 3.4 and the abuse liability axis of the AALF (Section S5).

**Figure S2. Peak change from baseline ($E_{max}$) in the Drug Effects Questionnaire Pleasurable composite, by dose and route.** Bars, group means ± SE; points, individual sessions. Brackets mark contrasts significant at $p < .05$.

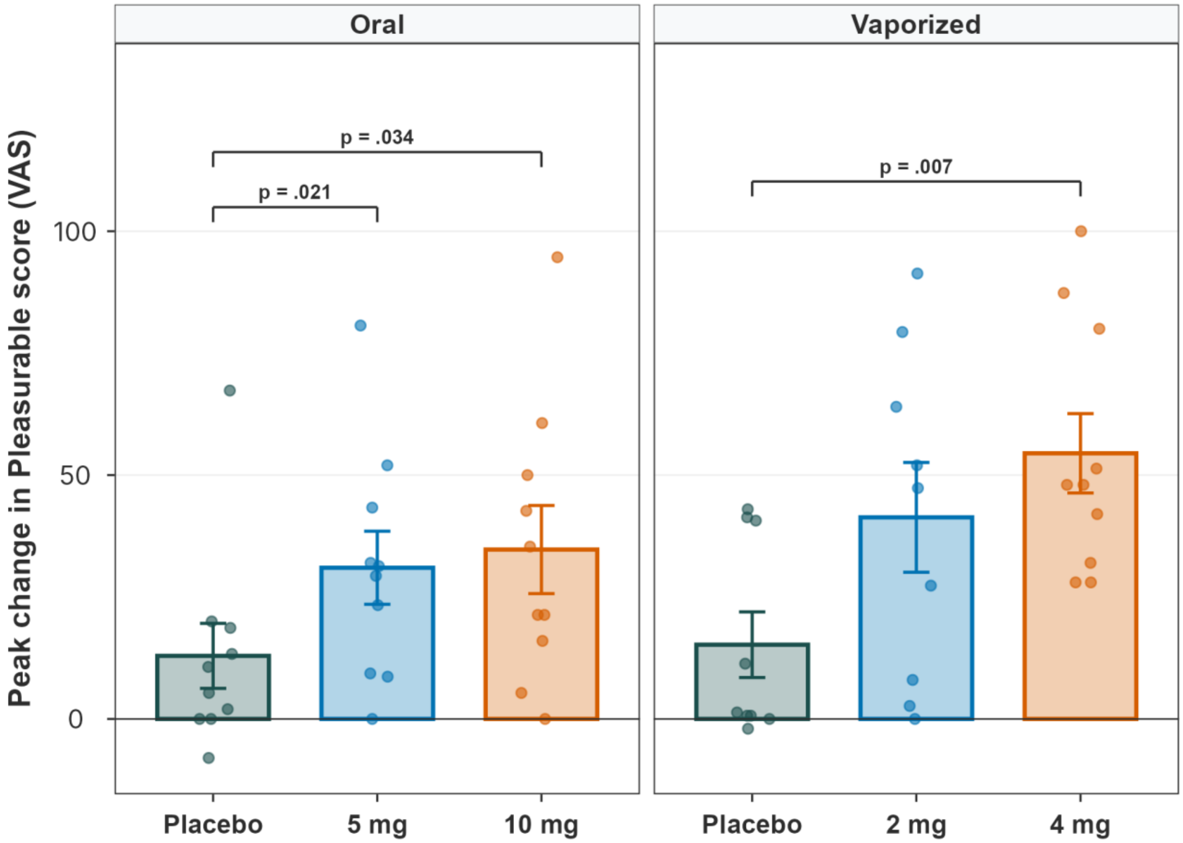

**S5. Analgesia-Abuse Liability Frontier**

A Pareto frontier identifies, among options (e.g., doses) judged by two outcomes (i.e., efficacy and safety), those that no other option dominates. It has guided dose selection where efficacy and toxicity rise together, as in tuberculosis [S1] and cancer combination therapy [S2]. Here, the Pareto frontier compares conditions on analgesia and abuse liability without combining them into one weighted score, so no exchange rate between the two is needed. We refer to our application as the Analgesia-Abuse Liability Frontier (AALF). To reduce the dimensionality of the QST data while capturing overall pain sensitivity, the analgesia axis was the pain sensitivity index, summarized as session AUC (main text Section 2.6). The abuse liability axis was the peak change in DEQ Pleasurable ($E_{max}$), the summary that FDA guidance recommends for primary analyses of abuse potential (main text Section 2.7).

Each active condition was placed at its model-estimated mean differences from placebo on the two outcomes (main text Section 2.8); bars show their 95% confidence intervals. Placebo, the reference, sits at the origin. Because both outcomes were measured in the same sessions, every condition could be placed on both axes at once. For each condition, we asked whether any other condition provided more analgesia at no greater abuse liability. A condition is dominated when another condition is better on at least one outcome and no worse on the other. Conditions that no other condition dominates form the AALF. Membership rests on the point estimates; the confidence intervals show how secure each position is. We rebuilt the frontier using reinforcement value (the Multiple-Choice Procedure crossover point) as the abuse liability axis and under all four combinations of session AUC and peak change for the two outcomes (Section S6). We also rebuilt it on each of the seven QST measures separately (Section S7). Frontiers were computed in R version 4.5.2.

Oral 5 mg was the only active condition on the AALF (**Figure S3A**). It produced the largest analgesic response and the lowest peak DEQ Pleasurable response of any active condition, raising the pain sensitivity index by 0.44 SD relative to placebo at a cost of 19.1 DEQ points. The other conditions produced less analgesia at a higher cost: oral 10 mg +0.05 SD at +22.8 points, vaporized 2 mg −0.19 SD at +21.0 points, and vaporized 4 mg −0.15 SD at +35.1 points. Oral 5 mg remained the only active condition on the frontier when abuse liability was indexed by reinforcement value (**Figure S3B**) and under all four combinations of session AUC and peak change (Section S6). On individual QST measures, it stayed on the frontier except on pressure pain tolerance and mechanical pain threshold, where vaporized 2 mg replaced it (Section S7; **Table S1**).

**Figure S3. Analgesia-Abuse Liability Frontier (AALF) for oral and vaporized Δ9-THC, abuse liability indexed by (A) subjective effects, the peak change in DEQ Pleasurable; and (B) reinforcement value, the Multiple-Choice Procedure crossover point.** Each active condition is placed at its model-estimated mean difference from placebo (main text **Table 2** and Section 3.4). Analgesia is on the vertical axis (standardized pain sensitivity index, session AUC; positive values denote analgesia) and abuse liability on the horizontal axis. Placebo, the reference, is the filled diamond at the origin. Filled circles denote active conditions on the frontier and open circles conditions for which another delivered more analgesia at no greater abuse liability cost. In A, the line from placebo to oral 5 mg is the frontier. In B there is no line: oral 5 mg produced more analgesia than placebo at a reinforcement value marginally below placebo's, so placebo is dominated and oral 5 mg alone forms the frontier. The axes through placebo are drawn as arrows pointing toward more analgesia (up) and more abuse liability (right), and each quadrant is tinted and labelled with its position relative to placebo: more analgesia with less abuse liability (upper left), more analgesia with more abuse liability (upper right), less analgesia with less abuse liability (lower left), and less analgesia with more abuse liability (lower right). Grey bars are 95% confidence intervals. Oral 5 mg was the only active condition on the frontier under both measures.

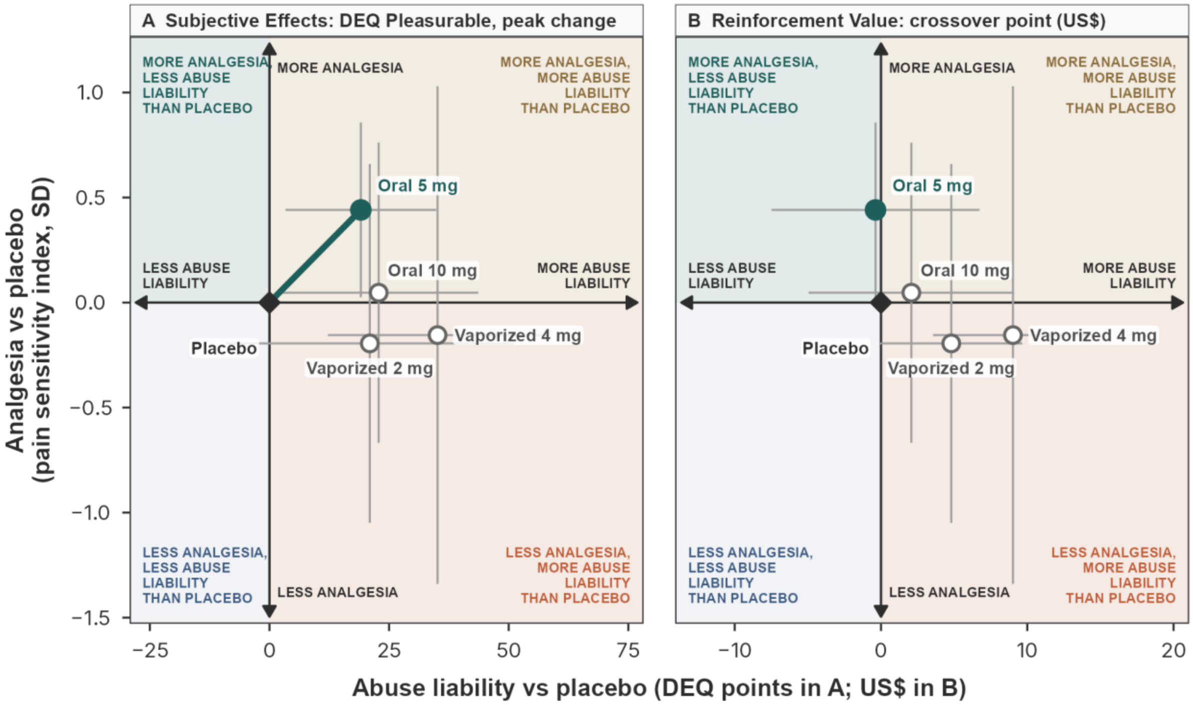

#### S6. Sensitivity of the Frontier to the Abuse Liability Measure and the Choice of Summary

The AALF (Section S5) summarizes analgesia as the session area under the change-from-baseline curve and abuse liability as the peak change in DEQ Pleasurable (primary analysis). Two sensitivity analyses varied these choices. When abuse liability was indexed by reinforcement value (the Multiple-Choice Procedure crossover point; **Figure S3B**), oral 5 mg was again the only active condition on the frontier. The frontier was also rebuilt under all four combinations of area and peak for the two outcomes. The outcome was identical in all four: oral 5 mg was the only active condition on the frontier, meaning that no other condition provided more analgesia at no greater abuse liability (**Figure S4**).

**Figure S4. The Analgesia-Abuse Liability Frontier (AALF) under all four combinations of session AUC and peak change for analgesia (rows) and abuse liability (columns).** Each active condition is placed at its model-estimated mean difference from placebo (main text **Table 2** and Section 3.4). Placebo, the reference, is the filled diamond at the origin; the axes through placebo are arrows pointing toward more analgesia (up) and more abuse liability (right), and each quadrant is tinted and labelled with its position relative to placebo. Filled circles denote active conditions on the frontier, and the line joins the conditions on the frontier; open circles denote conditions for which another delivered more analgesia at no greater abuse liability cost; grey bars are 95% confidence intervals.

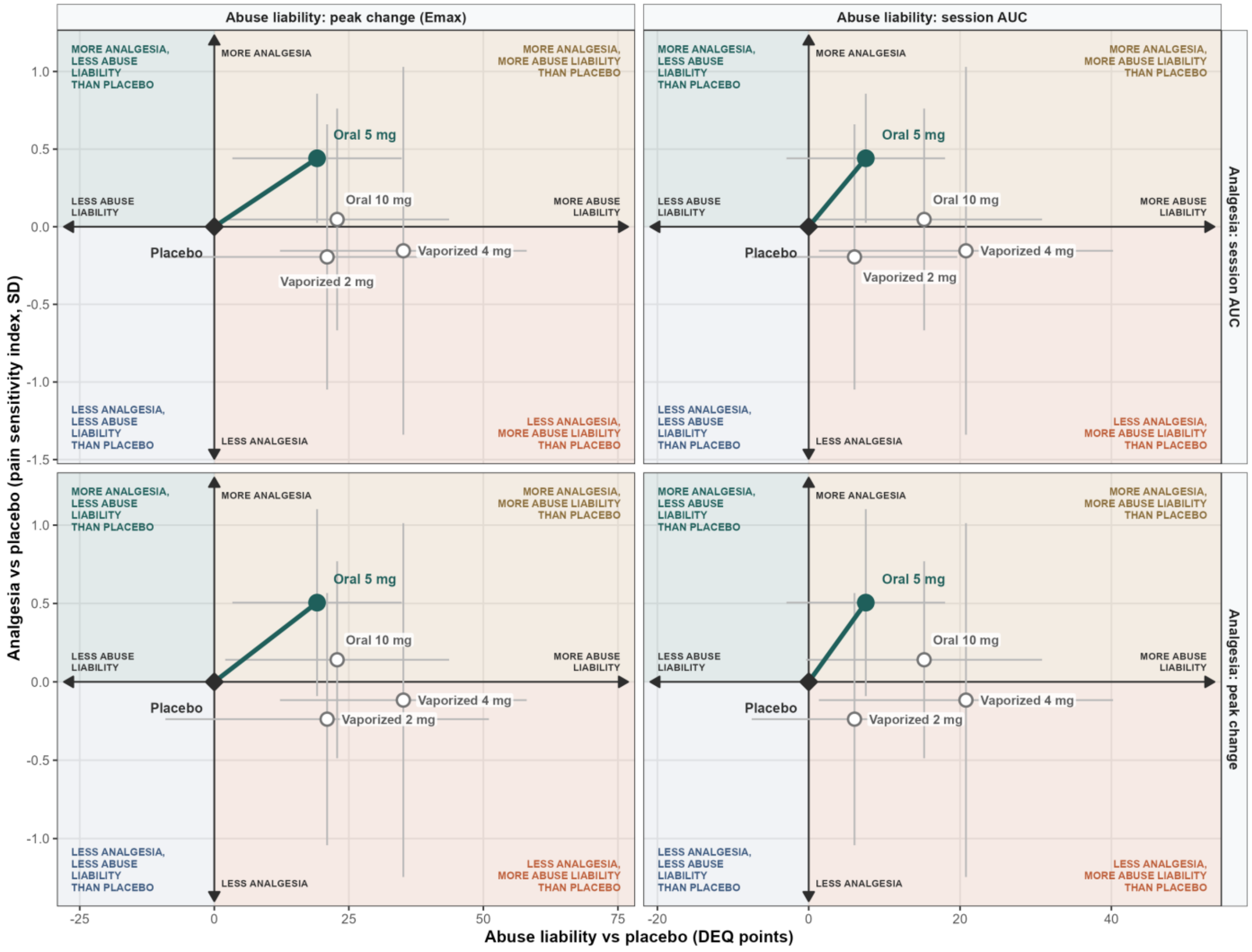

### S7. Sensitivity of the Frontier to Each QST Measure

The analgesia axis of the AALF is the pain sensitivity index, which combines the four thermal QST measures. The frontier was rebuilt on each of the seven QST measures alone, with every other element of the construction held constant (**Figure S5**; **Table S1**). Oral 5 mg was on the frontier for five of the seven measures and was the only active condition there for four: heat pain threshold, cold pain threshold, cold pain tolerance, and pressure pain threshold. On heat pain tolerance, vaporized 2 mg shared the frontier, producing more analgesia than oral 5 mg (0.60 vs 0.36 SD) at a higher abuse liability cost. On mechanical pain threshold and pressure pain tolerance, oral 5 mg produced less analgesia than placebo, and vaporized 2 mg was the only active condition on the frontier. The main analysis likewise found that vaporized 2 mg raised mechanical pain threshold above placebo (main text Section 3.2). Oral 10 mg and vaporized 4 mg were dominated on every measure.

**Figure S5. The Analgesia-Abuse Liability Frontier (AALF) rebuilt on each QST measure.** Analgesia is the session area under the change-from-baseline curve for that measure alone, in standard deviations; abuse liability is the peak change in DEQ Pleasurable. Each active condition is placed at its model-estimated mean difference from placebo. Placebo, the reference, is the filled diamond at the origin; the axes through placebo are arrows pointing toward more analgesia (up) and more abuse liability (right), and each quadrant is tinted and labelled with its position relative to placebo. Filled circles denote active conditions on the frontier, and the line joins the conditions on the frontier; open circles denote conditions for which another delivered more analgesia at no greater abuse liability cost; grey bars are 95% confidence intervals.

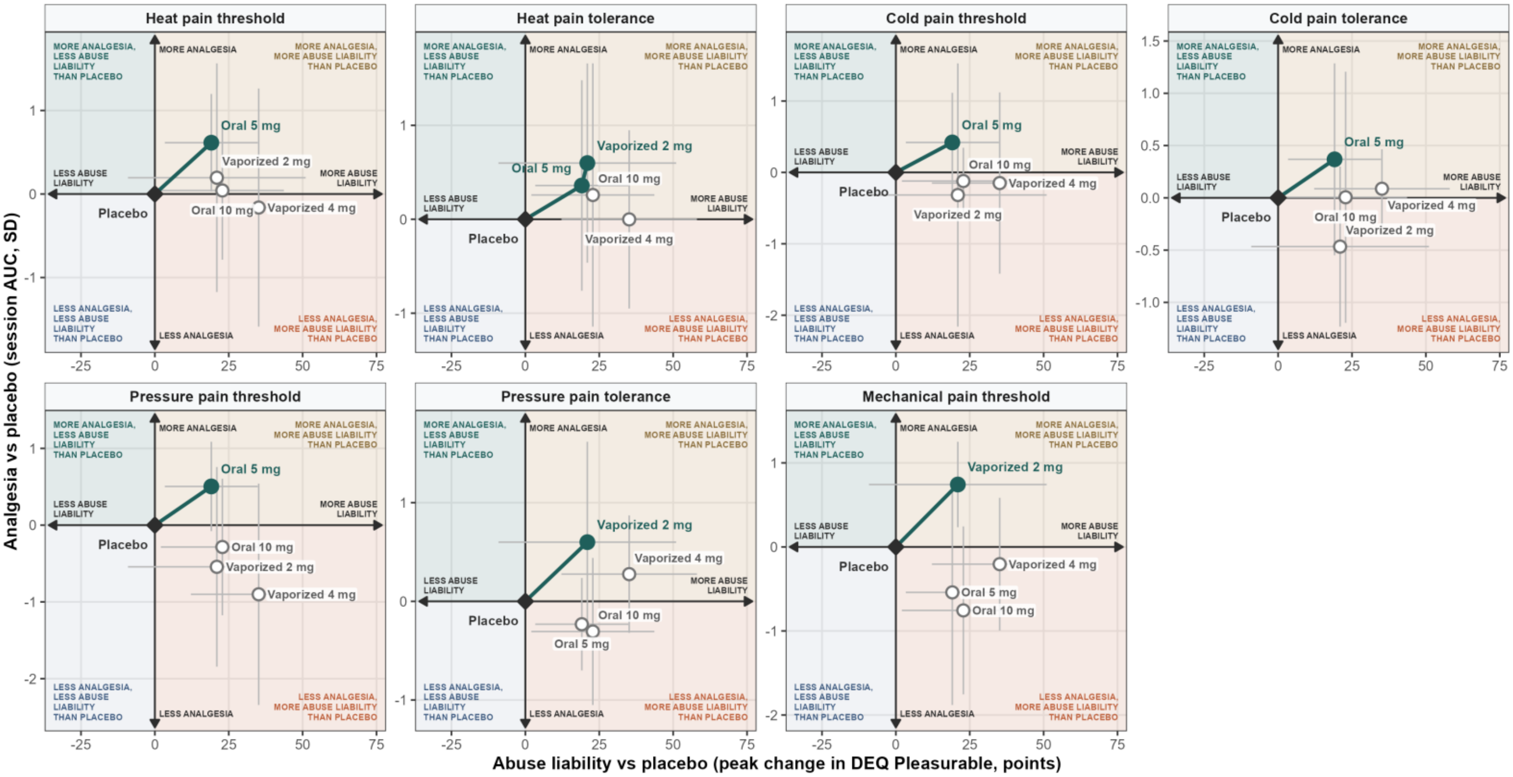

**Table S1. Active conditions on the frontier for each QST measure.** Placebo was on the frontier for every measure and is omitted. The last column is oral 5 mg's model-estimated difference from placebo on that measure alone, in SD units.

| Measure | Active conditions on the frontier | Oral 5 mg analgesia, SD (95% CI) |
|---|---|---|
| Heat pain threshold | Oral 5 mg | +0.61 (+0.03 to +1.20) |
| Heat pain tolerance | Oral 5 mg and vaporized 2 mg | +0.36 (−0.76 to +1.48) |
| Cold pain threshold | Oral 5 mg | +0.42 (−0.27 to +1.11) |
| Cold pain tolerance | Oral 5 mg | +0.37 (−0.55 to +1.29) |
| Pressure pain threshold | Oral 5 mg | +0.50 (−0.08 to +1.09) |
| Pressure pain tolerance | Vaporized 2 mg | −0.23 (−0.70 to +0.24) |
| Mechanical pain threshold | Vaporized 2 mg | −0.54 (−1.88 to +0.80) |

### S8. Exposure-Response

Each session's peak concentration and total exposure of the active combined analytes were related to analgesia and to both abuse liability measures with linear mixed models including a random intercept for participant, with routes pooled because route and peak exposure are confounded by design (**Figure S6**). Both abuse liability measures rose with exposure, the subjective measure more steeply with peak than with total exposure. No significant association between analgesia and either exposure metric was detected. Adding a quadratic term did not improve the analgesia models, so there was no sign of an inverted U. One oral 5 mg session whose exposure was recorded as zero, below the limit of quantification, was excluded from all exposure models. One vaporized 2 mg session without an estimable total exposure was excluded from the total-exposure models.

**Figure S6. Exposure-response for analgesia (A), subjective effects (B), and reinforcement value (C) against peak concentration (left) and total exposure (right) of the active combined analytes.** Points, individual sessions colored by route; lines, least-squares linear fits with 95% confidence bands; β and p from the mixed model, per standard deviation of exposure.

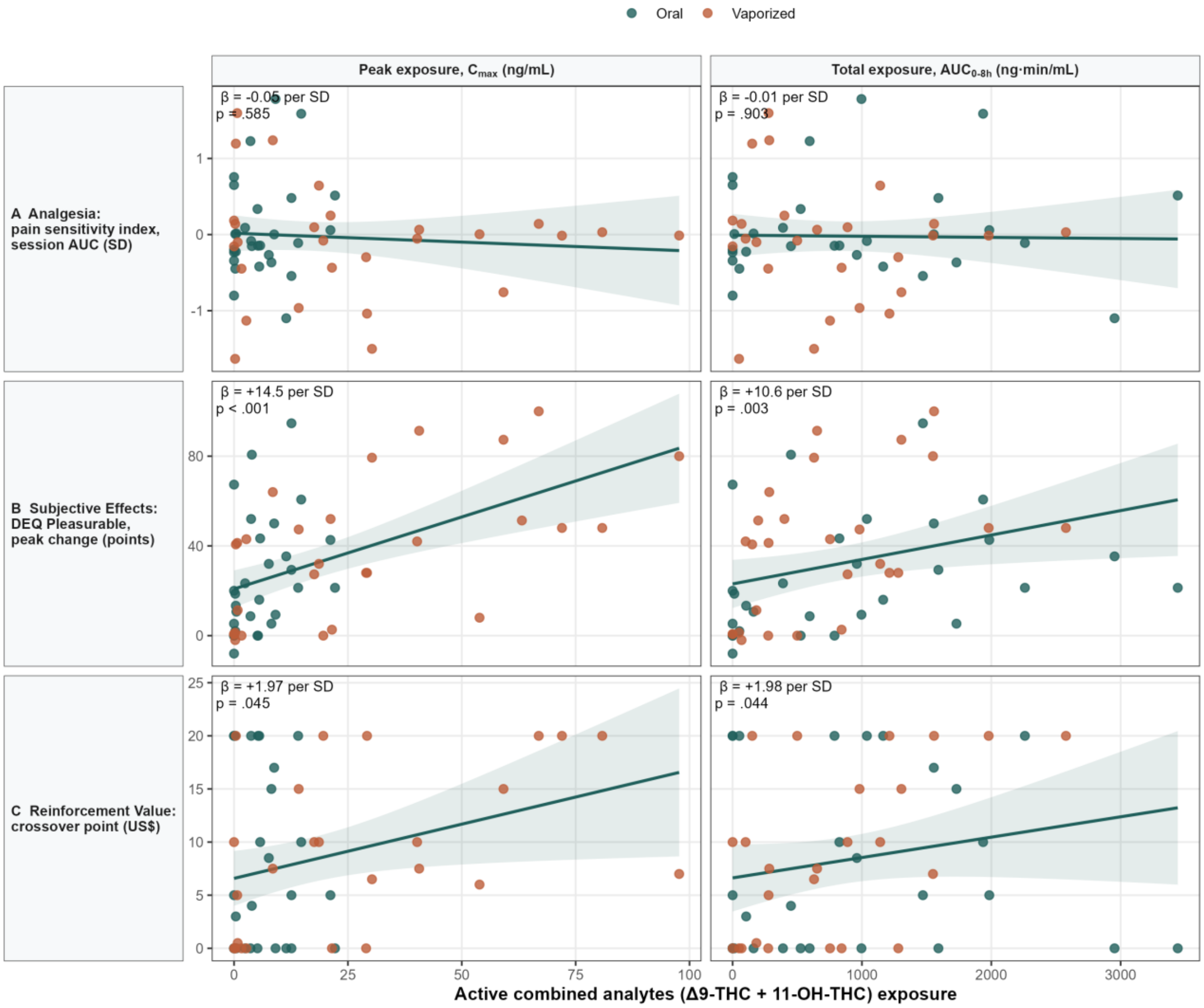

**S9. Time Courses of the Individual QST Measures**

Main text **Figure 3** shows the pain sensitivity index and DEQ Pleasurable over time; **Figures S7 and S8** show the individual QST measures. QST was sampled at 60, 120, 180, 240 and 360 min after oral dosing and at 5, 60, 120, 180 and 360 min after vaporized dosing. Each point averages 8 to 10 participants. To show the pain sensitivity index over time, we converted each thermal measure's change from baseline at each time point to the SD units of the index. We divided the change by the SD of that measure's session AUC across all sessions. We then multiplied it by the length of the session window (oral, 300 min; vaporized, 355 min). Finally, we averaged the four thermal measures. For sessions with no missing time point, each curve averaged over the session equals that session's pain sensitivity index plus a constant shared by all sessions.

**Figure S7. Time course of the four thermal QST measures, by route and dose.** Change from baseline in °C; the cold measures are sign-reversed so that positive values indicate less pain on every measure. Points, means; bars, standard errors.

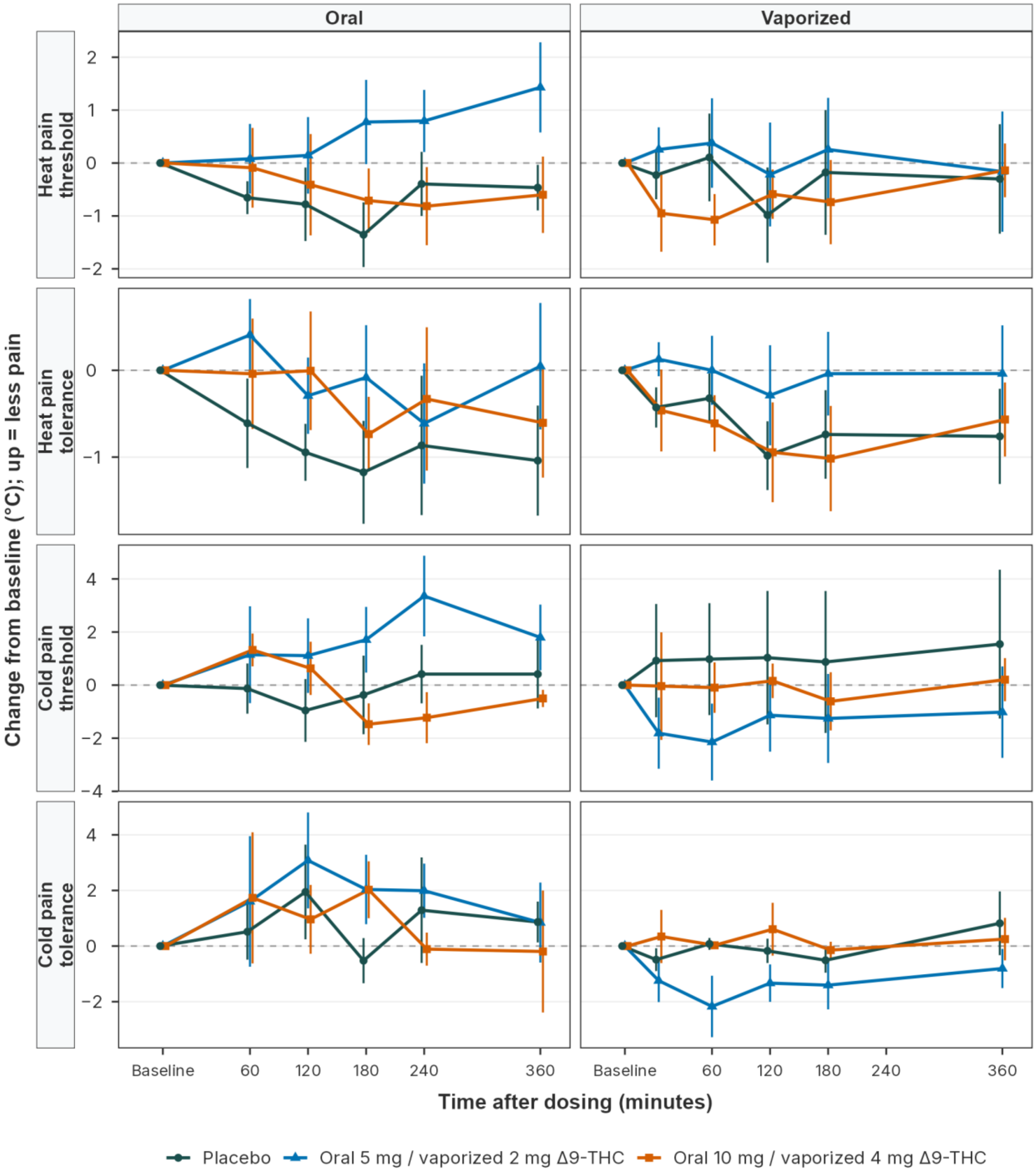

**Figure S8. Time course of pressure pain threshold and tolerance and of mechanical pain threshold, by route and dose.** Change from baseline in kPa (pressure) and mN (mechanical); positive values indicate less pain. Points, means; bars, standard errors.

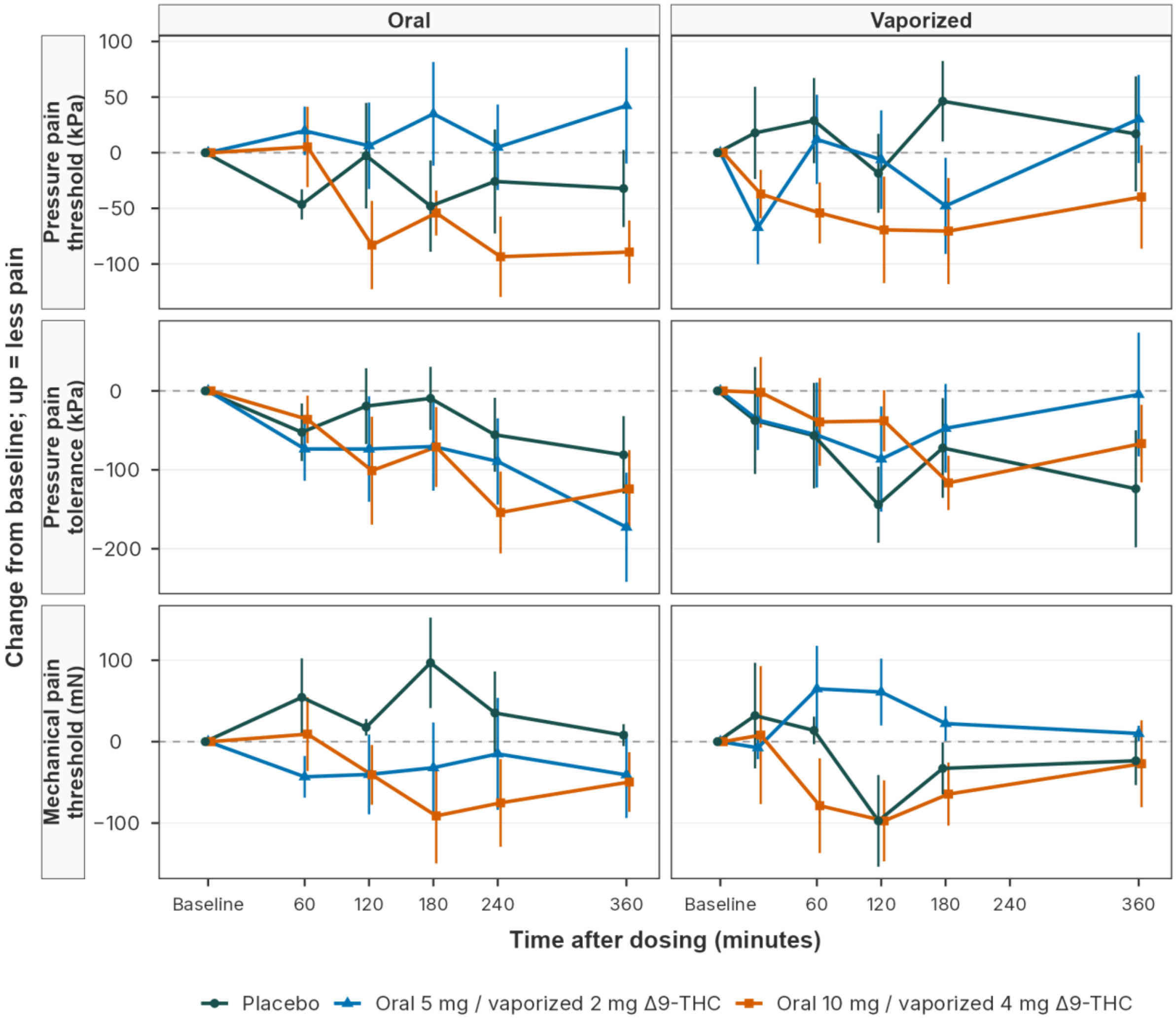
